# Functional connectivity of dorsolateral prefrontal cortex predicts cocaine relapse

**DOI:** 10.1101/2020.12.17.20245399

**Authors:** Tianye Zhai, Betty Jo Salmeron, Hong Gu, Bryon Adinoff, Elliot A. Stein, Yihong Yang

## Abstract

**Background:** Relapse is one of the most perplexing problems of addiction. The dorsolateral prefrontal cortex (DLPFC) is crucially involved in numerous cognitive and affective processes that are implicated in phenotypes of addiction, and is one of the most frequently reported brain regions with aberrant functionality in substance use disorders. However, the DLPFC is an anatomically large and functionally heterogeneous region, and the specific DLPFC-based circuits that contribute to drug relapse remain unknown.

**Methods:** We systematically investigated the relationship of cocaine relapse with 98 DLPFC functional circuits defined by evenly sampling the entire bilateral DLPFC in a cohort of cocaine dependent patients (n=43, 5F) following a psychosocial treatment intervention. A Cox regression model was utilized to predict relapse likelihood based on DLPFC functional connectivity strength.

**Results:** Functional connectivity from 3 of the 98 DLPFC loci, one on the left and two on the right hemisphere, significantly predicted cocaine relapse with an accuracy of 83.9%, 84.7% and 85.4%, respectively. Combining all three significantly improved prediction validity to 87.5%. Protective and risk circuits related to these DLPFC loci were identified that are known to support “bottom up” drive to use drug and “top down” control over behavior together with social emotional, learning and memory processing.

**Conclusion:** Three DLPFC-centric circuits were identified that predict relapse to cocaine use with high accuracy. These functionally distinct DLPFC-based circuits provide insights into the multiple roles played by the DLPFC in cognitive and affective functioning that affects treatment outcome. The identified DLPFC loci may serve as potential neuromodulation targets for addiction treatment and as clinically relevant biomarkers of its efficacy.

## Introduction

Drug addiction has serious negative impact on the individual, family, community and society at large, resulting in hundreds of billions of dollars in direct and indirect public costs annually attributed to crime, health care and loss of productivity (National Drug Intelligence Center (NDIC), 2011; Volkow *et al*., 2016). Preclinical and human research supports that drug addiction should be considered and treated as an acquired, highly relapsing (Dutra *et al*., 2008; Koob and Volkow, 2016), chronic brain disease (Volkow *et al*., 2016). Unfortunately, current addiction treatments remain relatively ineffective, with relapse rates post-treatment about 70%, and are further compounded by high dropout rates (approximately 35%) before treatment completion, even for treatment-seeking patients (Dutra *et al*., 2008).

These data reflect the core feature of the addiction phenotype: loss of control over drug use, which has been attributed to abnormalities in multiple cognitive and affective domains, including decision-making (Monterosso and Piray, 2012), inhibitory control (Zilverstand *et al*., 2018), craving (Garavan *et al*., 2000), memories (Hyman, 2005), and negative emotional regulation (Koob, 2008). These abnormalities are closely associated with dysfunctions in top-down executive control, which is mediated, at least in part, by regions within the dorsolateral prefrontal cortex (DLPFC) and their ‘downstream’ functional circuits (Dalley *et al*., 2011; Goldstein and Volkow, 2011).

As the core of the Executive Control Network (ECN) (Bressler and Menon, 2010; Seeley *et al*., 2007), the DLPFC is involved in many cognitive processes that are impaired in substance use disorders. DLPFC plays a pivotal role in decision-making, which is significantly dysregulated in substance use disorders (SUDs) (Clark and Robbins, 2002). As demonstrated via neuroeconomic models, the DLPFC contributes to intertemporal value-based decision-making in computing the value of long-term consequences of delayed vs. more immediate rewards (McClure *et al*., 2007). People with SUDs have a propensity for choosing more immediate smaller rewards over larger, delayed rewards when assessed via temporal discounting behavioral choices (Bickel *et al*., 2007; Monterosso and Piray, 2012). The DLPFC is implicated in risk avoidance. Using the Balloon Analogue Risk Task (Kohno *et al*., 2014), attenuated activity is seen in the right DLPFC that is accompanied by deficits in risk avoidance in methamphetamine dependent patients, compared to control subjects. The DLPFC is also involved in inhibition control, as demonstrated by activation during successful response inhibition using the Go/NoGo task (Garavan *et al*., 2006; Menon *et al*., 2001; Simmonds *et al*., 2008), and impaired response inhibition has been consistently demonstrated in SUD individuals accompanied with aberrant DLPFC activity (Goldstein and Volkow, 2011; Kaufman *et al*., 2003; Zilverstand *et al*., 2018). Cue-induced craving is another characteristic of SUD, where cues become associated with drugs by co-opting memory and the valuation systems, leading to drug wanting and seeking (Wimmer and Shohamy, 2012). The DLPFC has been implicated in cue-induced craving measured by both fMRI and PET (Bonson *et al*., 2002; Garavan *et al*., 2000; S. Grant *et al*., 1996; Maas *et al*., 1998; Wilson *et al*., 2004). In addition, the DLPFC is crucial in working memory (Barbey *et al*., 2013; Curtis and D’Esposito, 2003; Wesley and Bickel, 2014) that often show impairments in SUD (Goldstein and Volkow, 2011; Koob and Volkow, 2016). It has been demonstrated that working memory training can reduce delay discounting in stimulant addicted participants (Bickel *et al*., 2011). Decreased activity in the left DLPFC during an N-back working memory task was predictive of smoking relapse (Loughead *et al*., 2014), and poorer behavioral performance in a working memory embedded response inhibition task was accompanied by attenuated right DLPFC activation in cocaine addicted individuals (Hester and Garavan, 2004).

The DLPFC is also involved in several affective domains associated with addiction. Drug dependent individuals often suffer from a persistent negative emotional state similar to that seen in depressive disorders, which show high comorbidity with SUD (B. F. Grant *et al*., 2004; Koob and Volkow, 2010). In addition, the DLPFC is consistently shown to be hypoactive in major depressive disorder (MDD) (Siegle *et al*., 2007), and increased activity in this area is associated with recovery/remission of depressive symptoms (Brody *et al*., 2001; Koenigs and Grafman, 2009; Mayberg *et al*., 2000). Lesion studies causally link damage to the bilateral DLPFC with higher levels of depressive symptoms (Koenigs and Grafman, 2009). Moreover, the DLPFC is also implicated in explicit and implicit emotion regulation (Braunstein *et al*., 2017) which would be expected to be especially taxed during the preoccupation phase of addiction (Koob and Volkow, 2016).

Indeed, although multiple regions-of-interest (ROIs) within the DLPFC have been consistently implicated in brain functions related to SUD, the exact DLPFC loci and their associated circuits that underlie drug dependence and relapse remain unknown. Given the DLPFC’s central role in cognitive and affective processing and integration, and its implication in various neuropsychiatric diseases (Cole *et al*., 2014), which are often co-occurring with SUD (Conway *et al*., 2006), it would seem critical to better understand the functional circuitry that enables this large, heterogeneous brain region such a prominent role. As such, we interrogated the functional circuitry from individual ROIs covering the entire DLPFC and their relationship to post treatment relapse. Specifically, we utilized the Cox regression model combined with resting-state functional connectivity to produce a comprehensive map of DLPFC related circuits in a cohort of cocaine dependent individuals who were imaged after completing an inpatient psychosocial treatment regimen and subsequently followed post-treatment for 24 weeks (Fig. 1). We aimed to identify regions within the DLPFC and their corresponding functional circuits that relate to the likelihood of relapse to cocaine use.

**Fig. 1.**
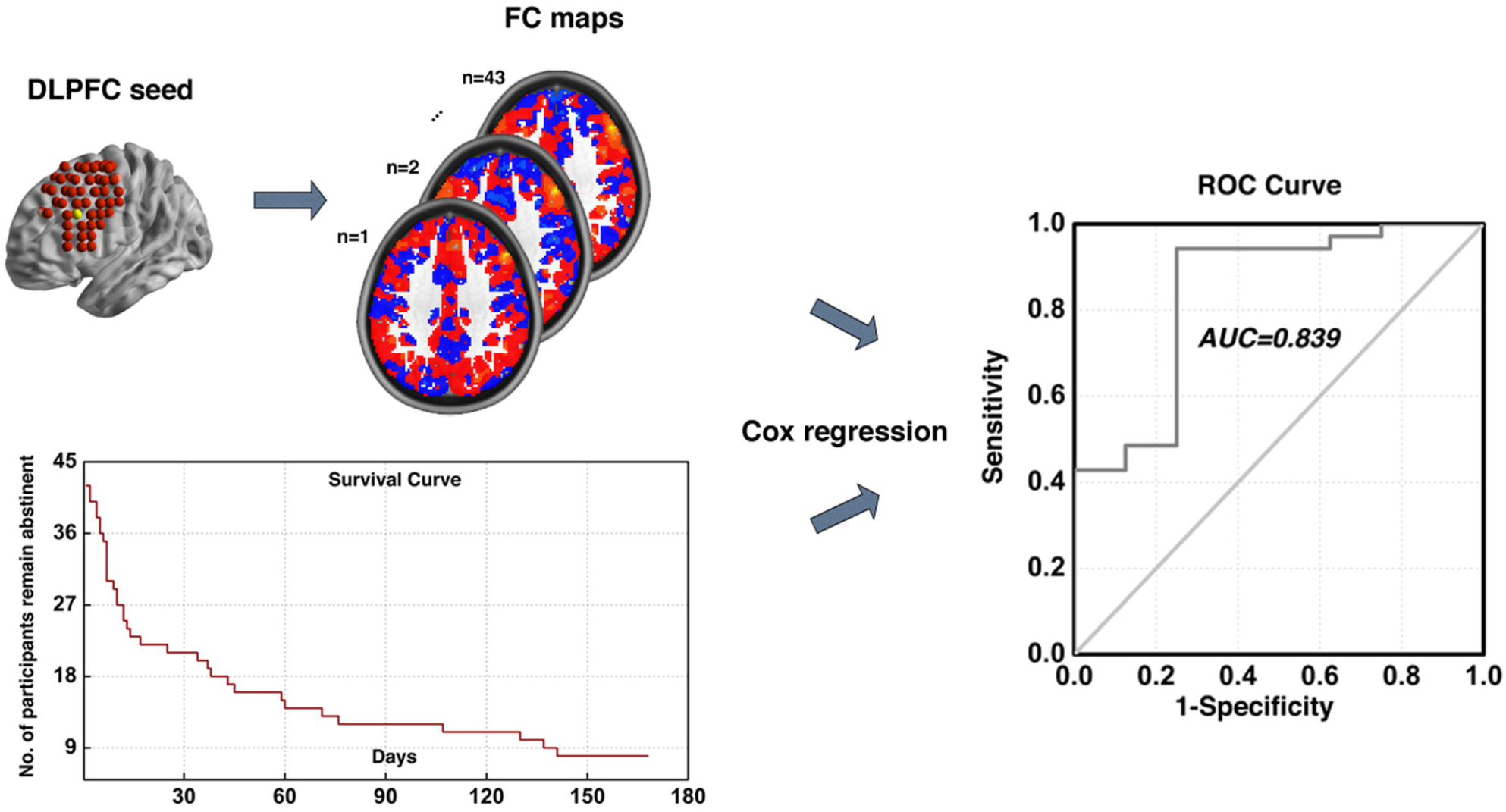
Illustration of analytical procedure. Illustration of our analytical procedure using one DLPFC locus as exemplar. First, the whole brain functional connectivity maps for each subject was calculated using the example DLPFC locus as seed. By combining these functional connectivity maps with the post-treatment information obtained during the follow-up period, we utilized the Cox regression based modeling to predict cocaine relapse. This procedure was conducted recursively for all 98 DLPFC sub-regions covering the entire bilateral DLPFC area.

## Materials and Methods

### Participants

Forty-five participants who completed treatment and follow-up from local residential treatment programs are included in current analysis. Study inclusion criteria included right-handedness, meeting criteria for cocaine dependence (DSM-IV), no history of major medical illness, estimated IQ over 70 based on the Wechsler Test of Adult Reading, not meeting criteria for any neurological or active Axis I disorder (other than substance dependence), and not on any psychotropic medications. The study was reviewed and approved by the Institutional Review Boards of the University of Texas Southwestern Medical Center and the Veterans Administration North Texas Health Care System. Written informed consent was obtained from each participant after the nature and possible consequences of the study were explained.

### Treatment and Assessment Procedures

The cocaine dependent participants were admitted to one of three treatment programs as soon as possible after their last reported use of cocaine. All three treatment programs utilized the Minnesota Model Psychosocial treatment approach (Cook, 1988). Demographic information and drug use history were collected during the first week of treatment. MRI scans were conducted during the final week of inpatient treatment. Urine drug screens (UDS) were conducted to verify abstinence.

Following discharge, participants were followed for up to 24 weeks or until relapse to stimulant use, whichever came first. During this period, two follow-up sessions occurred each week with one session over the phone and the other in-person. A structured interview assessing substance use since the previous visit (or since discharge from the treatment programs for the first visit) and a UDS (for the in-person session) were completed. Relapse was defined as any use of cocaine or amphetamine (either by self-report or by UDS) post-discharge or missing two consecutive appointments without contact. Date of relapse was recorded as the day of drug use or the day of the first missed appointment if lost to follow-up. Participants who failed to maintain abstinence were discharged from the study and excluded from further follow-up contact.

### MRI Acquisition

MRI scans were obtained using a Philips 3T scanner with an eight-channel radio-frequency coil (Philips Medical Systems, Best, The Netherlands). For each participant, six minutes of whole-brain blood-oxygen-level-dependent (BOLD) resting-state fMRI data were acquired in the axial plane parallel to the AC-PC line using a single-shot, echo-planar imaging sequence (TE=25ms, TR=1.7s, flip angle=70°, spatial resolution=3.25mm x 3.25mm x 3mm with no gap). Participants were instructed to keep their heads still and eyes open during the resting-state scan. A high-resolution anatomical T1-weighted image was also acquired from each participant using a 3D magnetization-prepared rapid gradient-echo sequence (TE=3.8ms, TR=8.2ms, flip angle=12°, spatial resolution=1mm x 1mm x 1mm).

### Data processing pipeline

The data processing pipeline consisted of 6 conceptual steps: 1) image preprocessing; 2) DLPFC functional connectivity calculation; 3) voxel-wise Cox regression analysis; 4) thresholding and generating composite indices; 5) Cox model fitting for brain-behavior relationship using the composite indices and model evaluation (ROC analysis); and 6) cross validation. Steps 3 to 6 were adapted and modified from the pipeline proposed by Shen, et al. (Shen *et al*., 2017), which provided a framework for modeling individual behaviors with whole brain functional connectivity. Supplementary Fig. 1 shows the schematic diagram of our analyses pipeline using one DLPFC locus as an exemplar, also see detailed description of steps in supplementary methods section.

### Data availability

Raw data generated in the current study contain personally identifiable information that could compromise the privacy of research participants by sharing publicly. Derived data supporting the findings of this study are available from the corresponding author contingency of institutional approval, upon reasonable request.

## Results

### Demographic and clinical characterization

Of the 45 participants, 43 remained in the final analysis following removal of 2 participants with excessive head motion. This cohort included 5 females and 38 males, with a mean (SD) age of 43.4 (7.2), years of education of 12.5 (2.1), years of cocaine use of 8.3 (5.2), and cigarette per day (CPD) of 11.3 (10.3). As shown in the survival curve in Fig. 1 (lower left corner), the number of participants who remained abstinent dropped rapidly during the first 30 days, and by the end of the 168-day follow-up period, 35 out of 43 participants had relapsed.

### Heterogeneity in predicting relapse across multiple DLPFC ROIs

The investigation across the 98 ROIs covering the surface area of both the left and right DLPFC yielded prediction models with AUC values of ROC curves ranging from 0.343 to 0.854. Followed by a conservative correction (Bonferroni) for multiple comparisons, three of the 98 DLPFC ROIs (one on the left and two on the right side) significantly predicted cocaine relapse with their corresponding functional circuits (*p* < 5.1×10^−4^, 0.05/98). The ROI on the left side of DLPFC (ROC curve showing AUC of 0.839) was located at MNI coordinates [-48, 30, 34] (Fig. 2A, B). We define this locus as ‘predictive ROI-1’ hereafter. Two ROIs on the right side that showed significant prediction accuracy (ROC curves showing AUC value of 0.847 and 0.854) were located at MNI coordinates [32, 46, 34] and [32, 30, 50] (Fig. 3A, B, and Fig. 4A, B). We define these two ROIs as ‘predictive ROI-2’ and ‘predictive ROI-3’, respectively.

**Fig. 2.**
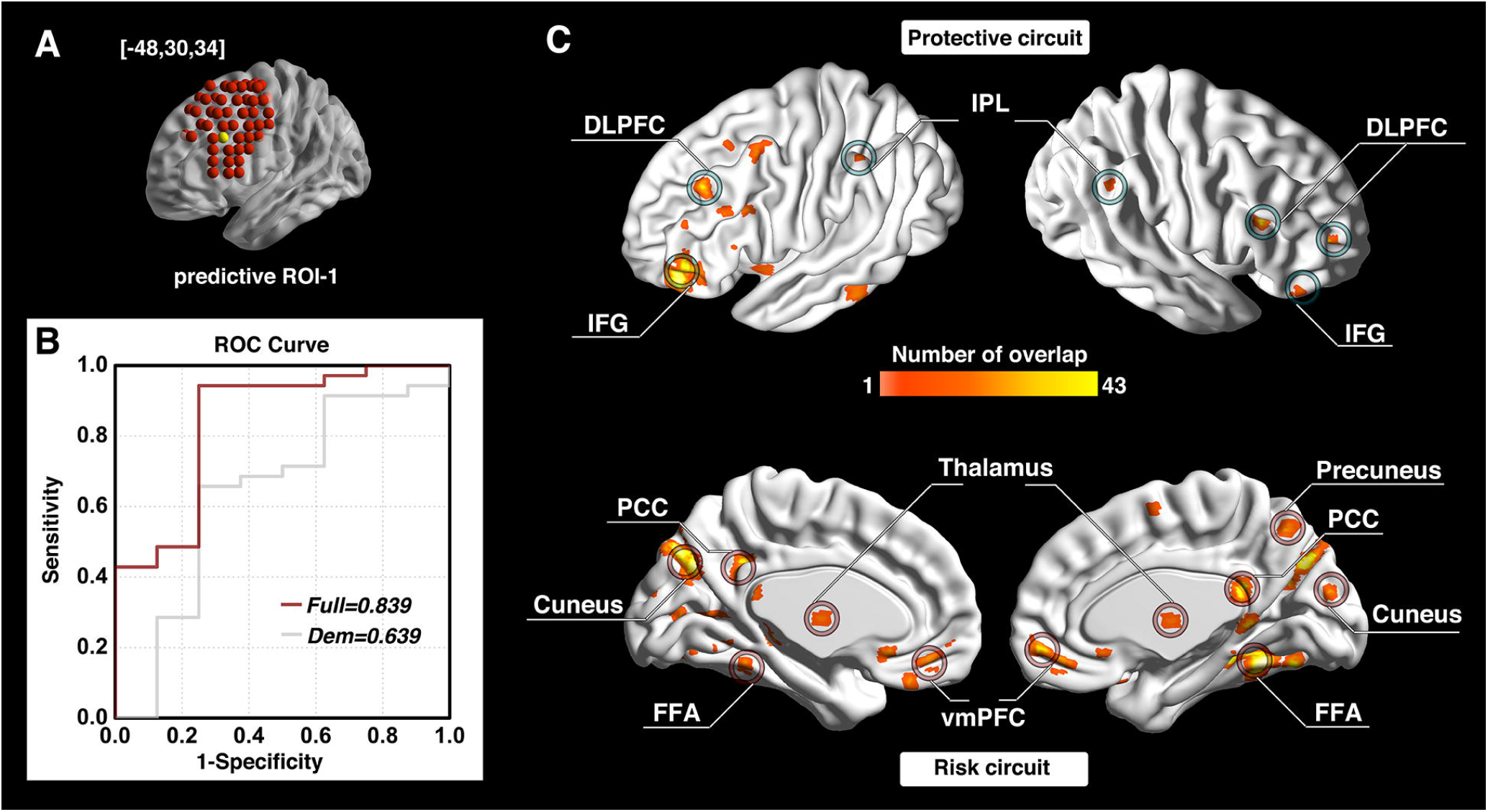
Prediction accuracy and functional circuits that contribute to cocaine relapse prediction (predictive ROI-1) Relapse prediction based on predictive ROI-1 [-48, 30, 34] **(yellow ball in A)** yielded AUC value of 0.839 **(B)**. Two functional circuits were identified that contributed to cocaine relapse: The protective circuit includes bilateral DLPFC, inferior frontal gyrus (IFG), and inferior parietal lobule (IPL) **(C, upper)**; the risk circuit includes the ventromedial prefrontal cortex (vmPFC), thalamus and the posterior cingulate cortex (PCC)/precuneus (default mode network regions), as well as cortical regions, including visual and motor regions **(C, lower)**. *Abbreviations:* ROI, region of interest; ROC, receiver operating characteristic; Dem, demographic model (prediction model only use demographic information); DLPFC, dorsolateral prefrontal cortex; IFG, inferior frontal gyrus; IPL, inferior parietal lobule; PCC, posterior cingulate cortex; vmPFC, ventromedial prefrontal cortex; FFA, fusiform face area.

**Fig. 3.**
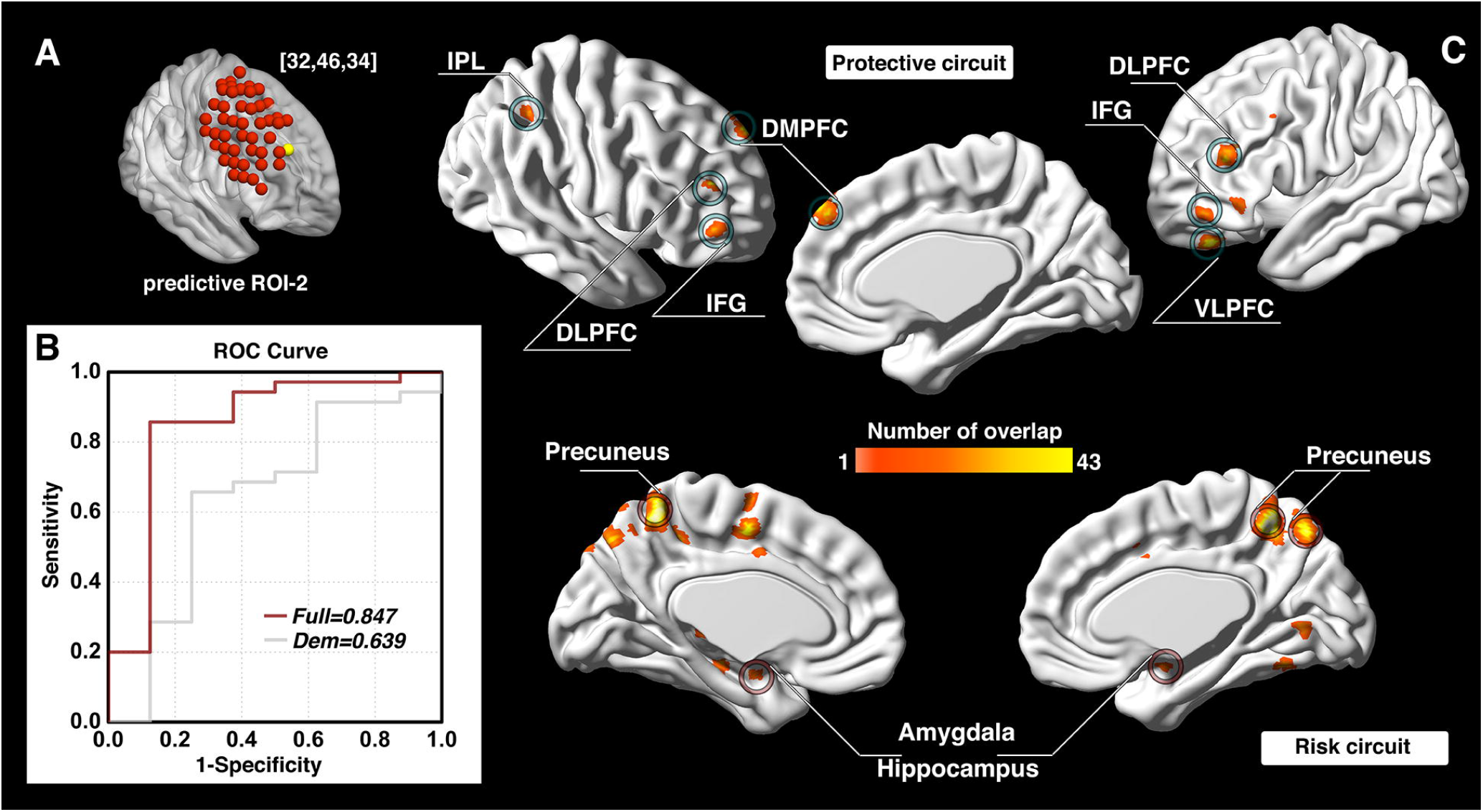
Prediction accuracy and functional circuits that contribute to cocaine relapse prediction (predictive ROI-2) Relapse prediction based on predictive ROI-2 [32, 46, 34] **(yellow ball in A)** yielded AUC value of 0.847 **(B)**. Two functional circuits contributed to cocaine relapse: The protective circuit mainly includes bilateral DLPFC, IFG, right IPL, DMPFC, and left VLPFC **(C, upper)**; the risk circuit mainly includes bilateral precuneus and amygdala/hippocampus **(C, lower)**. *Abbreviations:* ROI, region of interest; ROC, receiver operating characteristic; Dem, demographic model (prediction model only use demographic information); DLPFC, dorsolateral prefrontal cortex; IFG, inferior frontal gyrus; IPL, inferior parietal lobule; DMPFC, dorsomedial prefrontal cortex; VLPFC, ventrolateral prefrontal cortex.

**Fig. 4.**
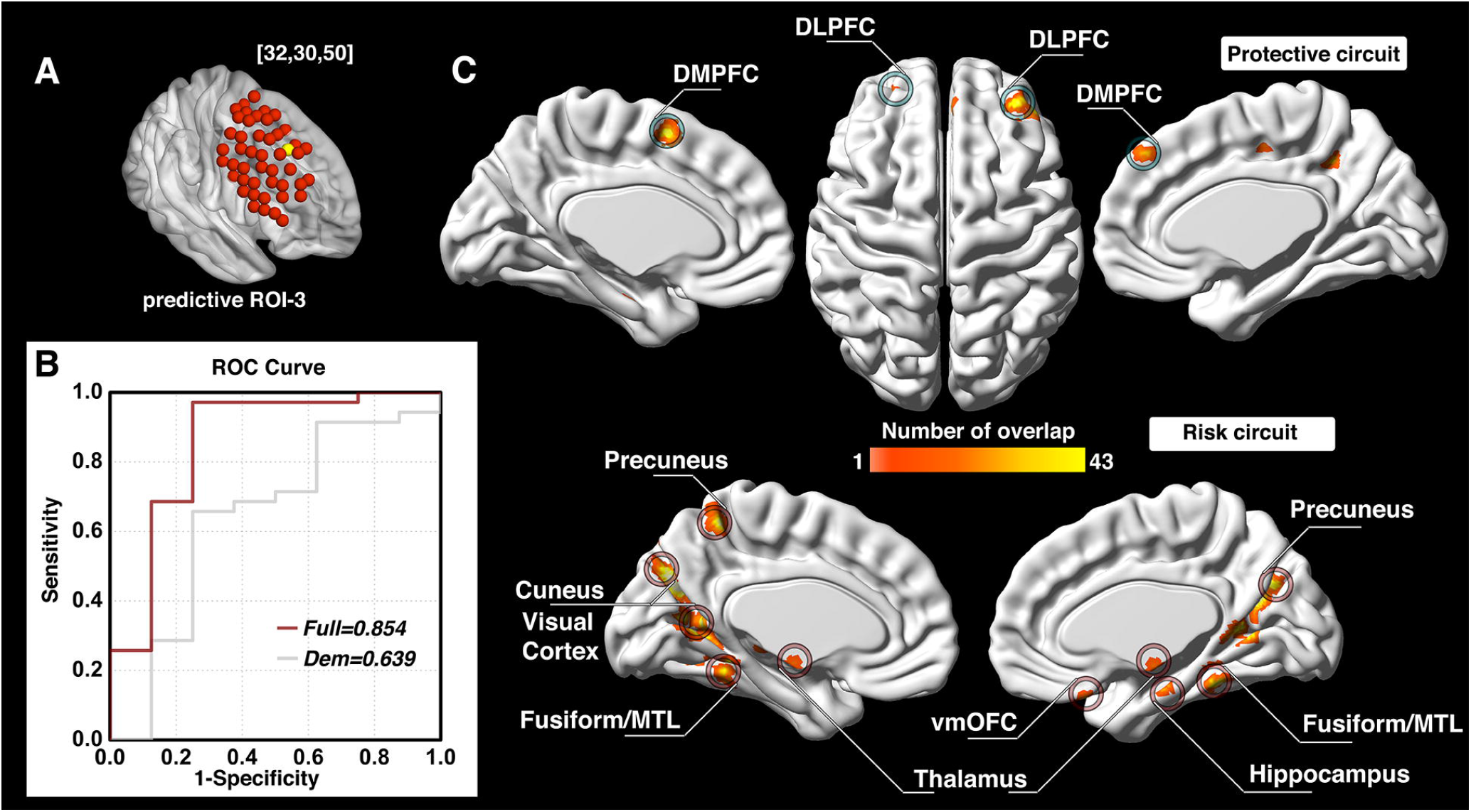
Prediction accuracy and functional circuits that contribute to cocaine relapse prediction (predictive ROI-3) Relapse prediction based on predictive ROI-3 [32, 30, 50] **(yellow ball in A)** yielded AUC value of 0.854 **(B)**. Two functional circuits contributed to cocaine relapse: The protective circuit mainly includes bilateral DLPFC and DMPFC **(C, upper)**; the risk circuit included the bilateral cuneus/visual cortex, vmOFC, thalamus, fusiform gyrus, and the left precuneus **(C, lower)**. *Abbreviations:* ROI, region of interest; ROC, receiver operating characteristic; Dem, demographic model (prediction model only use demographic information); DLPFC, dorsolateral prefrontal cortex; DMPFC, dorsomedial prefrontal cortex; vmOFC, ventromedial orbitofrontal cortex; MTL, medial temporal lobe.

### Neural circuits related to the three predictive ROIs confer protection against and risk for cocaine relapse

For each of the predictive loci, the nature of the Cox regression based survival analysis allow us to identify two sets of DLPFC functional circuits that uniquely contributed to cocaine relapse: a set of ‘protective’ circuits, which consisted of voxels whose connectivity with the DLPFC ROI correlated negatively with relapse likelihood (the stronger functional connectivity, the less probability of relapse within the follow-up period); and a set of ‘risk’ circuits which consisted of voxels whose connectivity with the DLPFC ROI correlated positively with relapse likelihood (the stronger functional connectivity, the higher probability of relapse within the follow-up period). For predictive ROI-1 (Fig. 2C), the protective circuits comprised components of the canonical executive control network (ECN) (Raichle, 2015), including the bilateral DLPFC, inferior parietal lobule (IPL), and inferior frontal gyrus (IFG); while the risk circuits comprised DLPFC connections to the canonical default mode network (DMN) (Raichle, 2015), including the bilateral ventromedial prefrontal cortex (vmPFC), posterior cingulate cortex (PCC)/precuneus, thalamus, along with visual and motor cortices, and the fusiform face area (FFA). For predictive ROI-2 (Fig. 3C), the protective circuits include the bilateral DLPFC, IFG, right IPL, DMPFC, and left ventrolateral prefrontal cortex (VLPFC), while the risk circuits comprised primarily the bilateral precuneus and amygdala/hippocampus. For predictive ROI-3 (Fig. 4C), the protective circuits included the bilateral DLPFC and DMPFC, while the risk circuits included the bilateral cuneus/visual cortex, ventromedial orbitofrontal cortex (vmOFC), thalamus, fusiform gyrus, and left precuneus. (See Supplementary Table 1 for a detailed list of circuit regions).

### Prediction model combining circuits from all 3 predictive ROIs

To determine if prediction models from these three DLPFC loci explained variance in relapse in an overlapped or independent fashion, we combined the indices for the protective and the risk circuits from all three loci and built a *combined model*. As shown in Fig. 5A, B, the *combined model* (predictive ROI-1+ROI-2+ROI-3) yielded an AUC of 0.875, demonstrating a significant improvement over the predictive ability of each of the three predictive ROIs alone, based on log-likelihood testing (Fig. 5C, *p*<0.001 for all three comparisons).

**Fig. 5.**
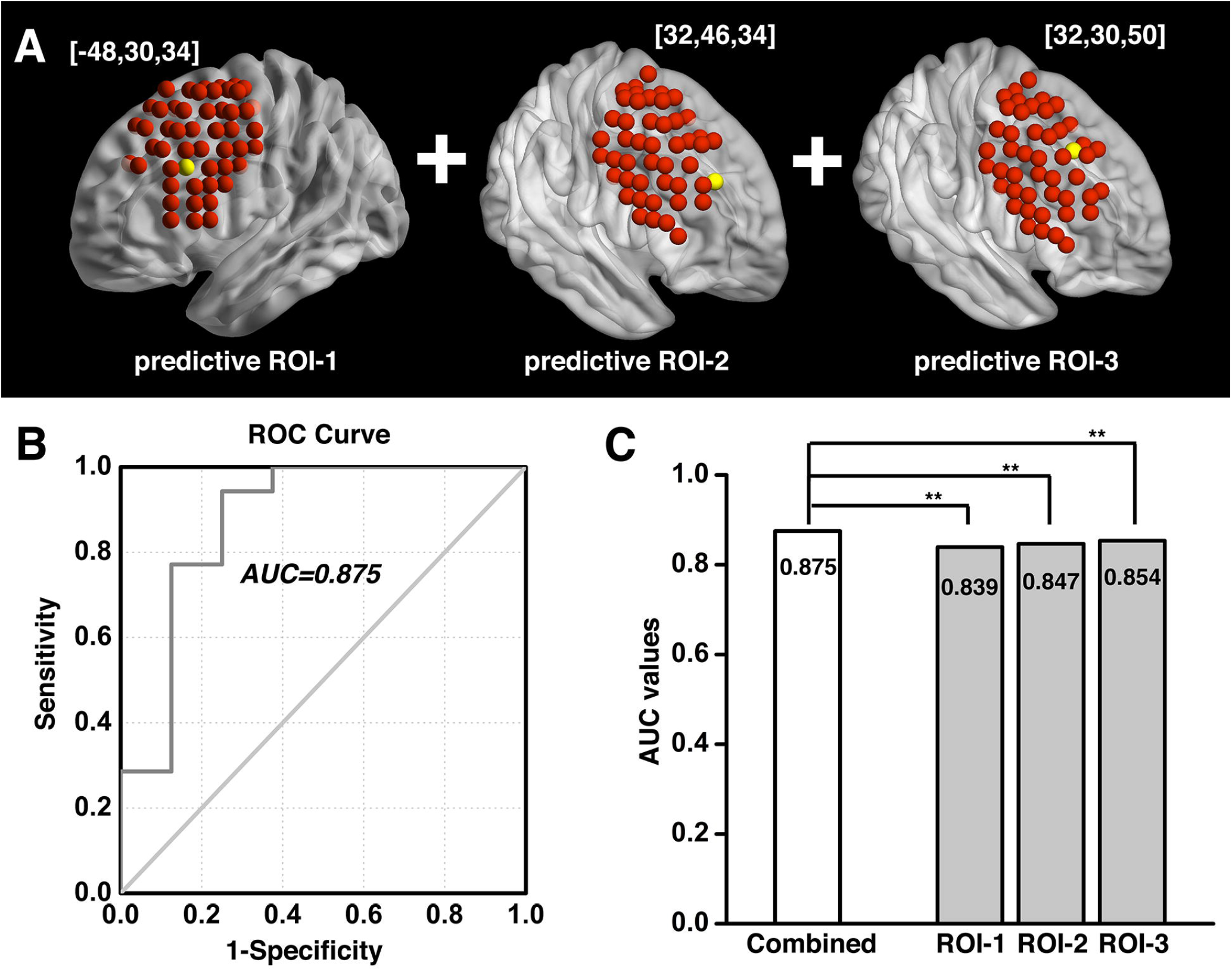
Combined prediction model based on predictive ROI-1, -2, and -3. When combining the protective and risk circuits from all three predictive ROIs **(A)**, the prediction model yielded an AUC of 0.875 **(B)**, showing significant improvement over each of the three individual predictive ROIs taken alone, based on log-likelihood testing **(C)**. **: *p*<0.001. *Abbreviations:* ROI, region of interest; ROC, receiver operating characteristic; AUC, Area-Under-Curve.

## Discussion

Using resting-state functional connectivity and a Cox regression-based prediction model, we systematically investigated the relationship between functional connectivity from multiple DLPFC ROIs and treatment outcome in a cohort of cocaine dependent individuals who had completed an inpatient, psychosocial treatment intervention. From the 98 ROIs covering the entire bilateral DLPFC surface area, we identified 3 sub-regions (predictive ROI-1, 2, and 3) that demonstrated predictive validity of cocaine relapse within the 24-week study follow-up period. Moreover, the Cox regression allowed us to identify two sets of functional circuits for each of these DLPFC ROIs, one related to risk for and the other related to protection against relapse (referred to as ‘risk circuit’ and ‘protective circuit’ hereafter). These circuits have previously been associated with DLPFC functions that are consistent with the cocaine dependence phenotype, as discussed below.

### The protective and risk circuits from predictive ROI-1

The protective circuit identified from the left hemisphere DLPFC ROI (predictive ROI-1; Fig. 2A) include the bilateral DLPFC, IPL, and IFG, all of which are components of the canonical ECN, which has consistently been implicated in top-down executive control, response inhibition and performance of attentionally demanding cognitive tasks (Menon, 2011; Moeller *et al*., 2016; Smith *et al*., 2014). Reduced functional connectivity strength within ECN regions has previously been reported in cocaine dependent individuals (Kelly *et al*., 2011; McHugh *et al*., 2017). This identified protective circuit has also been termed the “δ-network” within a neuroeconomic based decision-making context, and serves as a “control” network that takes long-term considerations into account when facing alternative valuation choices (McClure *et al*., 2007; Monterosso and Piray, 2012). Consistent with these previous findings, this identified protective circuit would suggest a stronger “control” network lowers the likelihood of relapse (i.e., increase abstinence likelihood). Indeed, one of the key symptoms of substance dependence, impaired response inhibition, has been associated with abnormalities (generally reduced task-induced activation compared to healthy control individuals) in these same ECN regions (i.e. DLPFC, IFG and IPL) (Cieslik *et al*., 2015; Moeller *et al*., 2016; Smith *et al*., 2014). Longitudinal studies further show that impaired response inhibition along with its underlying neural correlates is associated with not only the onset of substance use in adolescents with little previous substance use experience, but also relapse in dependent individuals attempting to quit (Moeller *et al*., 2016; Norman *et al*., 2011).

In contrast, the risk circuits associated with predictive ROI-1 interconnect the DLPFC with the vmPFC, PCC/precuneus, visual and motor cortex, thalamus and fusiform face area (Fig. 2C). Critically, these regions largely overlap with the well-established Default Mode Network (DMN), characterized by higher activity during internally-oriented thoughts and suppressed during performance of cognitively-demanding tasks (Raichle and Snyder, 2007). As suppression of DMN activity is thought to be required for optimal performance of goal directed behavior (Sutherland *et al*., 2012), stronger DMN functional connectivity with this DLPFC locus may underlie the enhanced drug craving and impaired goal-directed behavior seen in SUD individuals and may reflect less capacity to disengage DMN and in turn, enhance ECN to promote optimal performance, as suggested by large-scale network models of neuropsychiatric diseases (Menon, 2011; Sutherland *et al*., 2012). Acute and chronic stress and craving are known risk factors for SUD relapse (Seo *et al*., 2013; Shaham *et al*., 2003). For example, stress induced alcohol craving correlates with increased vmPFC and PCC/precuneus activity, which can serve as a predictor of both shorter time to relapse and heavy drinking in alcohol use disorder (AUD) patients (Seo *et al*., 2013).

This identified risk circuit of predictive ROI-1 is also termed the “β-network” in a neuroeconomic based decision-making context, and is thought to serve as a “drive” network that primarily mediates immediate/short-term reward (McClure *et al*., 2007; Sharp *et al*., 2012). Our results are consistent with this model by relating stronger functional connectivity between the DLPFC and these β-network regions to higher relapse likelihood. Given that steep discounting for delayed reward and the dysregulated interaction between these two networks is a known characteristic of SUD (Bickel *et al*., 2007; Monterosso and Piray, 2012), that this ROI and its related circuits predicted relapse with a high level of accuracy is supportive of this framework. Enhanced β-network functional connectivity and/or blunted δ-network functional connectivity (Zhai *et al*., 2015) has also been associated with other SUD dysregulated behaviors, including impulsivity and compulsive drug taking (Hu *et al*., 2015; Zhai *et al*., 2015). Importantly, the protective and risk related DLPFC circuits identified herein also suggest that the *balance* of distinct DLPFC based circuits related to immediate desires versus long-term planning is an important factor in individual treatment outcomes.

### The protective and risk circuits from predictive ROI-2 and ROI-3

The other two predictive DLPFC ROIs (ROI-2, Fig. 3A; and ROI-3, Fig. 4A) are located on the right hemisphere. The protective circuits associated with ROI-2 mainly include the bilateral DLPFC, IFG, right IPL, DMPFC, and left VLPFC, while the protective circuits associated with ROI-3 mainly consist of the bilateral DLPFC and DMPFC. While the VLPFC is typically associated with response inhibition (Sturm *et al*., 2016), the DMPFC is crucially involved in social-emotional processes such as negative emotions and social judgments (Etkin *et al*., 2011; Ferrari *et al*., 2016). It also plays an important role in stress-induced reinstatement of drug-seeking behavior (Feltenstein and See, 2008). Negative emotional states caused by drug withdrawal are a key causation of addiction relapse (Koob and Volkow, 2010) as well as one of the major causes of SUD establishment, maintenance and relapse (Koob and Volkow, 2016). Interestingly, the DMPFC has been implicated in Theory of Mind (ToM) (Isoda and Noritake, 2013), defined as the ability to attribute mental states to oneself and especially to others (Premack and Woodruff, 1978), a crucial feature in everyday life and social interaction that is impaired in SUD (Sanvicente-Vieira *et al*., 2017). It is well documented that cocaine dependent individuals show deficits in social-emotional processing (Geng *et al*., 2017; Preller *et al*., 2014). Our finding that the functional connectivity between DLPFC and DMPFC and VLPFC conveys protection against relapse may speak to the importance of the ability of the ECN, and DLPFC in particular, to regulate negative emotions and broader social-emotional processing necessary for successful outcomes in CUD treatment. The inclusion of the IFG, IPL, and the contralateral DLPFC in this protective circuit once again highlights the importance of a well-connected ECN in maintaining successful abstinence.

In contrast, the risk circuit associated with ROI-2 comprised primarily bilateral precuneus and the amygdala/hippocampus, while the risk circuit associated with ROI-3 included the bilateral cuneus/visual cortex, vmOFC, thalamus, fusiform gyrus and left precuneus. SUD has been characterized as a disease of dysregulated learning and memory, whereby maladaptive long-term memory formation pathologically usurps normal homeostatic processes to support continued drug use (Hyman, 2005). Reward-related learning involves multiple memory systems, including reinforcement learning via the mesocorticolimbic (MCL) pathway, and declarative memory systems of the medial temporal lobes (MTLs) and the precuneus (Adcock *et al*., 2006; Delgado and Dickerson, 2012; Wimmer and Shohamy, 2012). The MCL dopaminergic system, hippocampus and amygdala are progressively recruited in different phases of learning (Lin *et al*., 2019; Lüscher, 2016). SUD is also characterized by intense drug cravings that lead to stressful, aversive feelings, which may also lead to negative reinforcement, drug taking and relapse (Koob, 2015; Lüscher, 2016). Our finding that the functional connectivity between DLFPC and the MTL–precuneus declarative memory system, along with the circuit between DLPFC and the MCL system (thalamus and vmOFC) may serve as risk factors to relapse is likely a reflection of the negative impact of both long term alterations in declarative memory systems and the aberrant conventional reward-based, non-declarative memory systems dependent on ECN control of behavior in SUD.

The identification of distinct risk and protective circuits from the three predictive ROIs also speaks to the heterogeneous phenotype of the disorder and may help characterize the well-known individual differences in cocaine dependence. For example, cocaine dependent individuals may differ in the relative balance of very strong drive coupled with relatively normal control ability while others may have moderate drive levels but impaired cognitive control. For others, ability to manage social and emotional challenges may contribute more strongly to their substance dependence than executive function, per se. Such hypothetical distinct endophenotypes are otherwise difficult to distinguish during standard pre-treatment assessments. Identifying individual CUD subtypes based on circuits related to risk and protection may allow assessment of which circuits are most impaired in a given individual, leading to more individualized, and hopefully more efficacious interventions.

### Implications for Neuromodulation-based treatment

Taken together, the three treatment outcome predictive DLPFC ROIs may have important implications for potential TMS neuromodulation interventions. The predictive ROI-1 that showed high relapse predictive validity (AUC of 0.839) along with its functional circuits (both protective- and risk-circuit) is located within the left hemisphere DLPFC. Intriguingly, an open label pilot study in which rTMS significantly reduced cocaine relapse compared to a group receiving pharmacological interventions (Terraneo *et al*., 2016) employed a left DLPFC stimulation target that is only 2.83 mm proximal to this ROI. The consistency between our predictive validity and a real world TMS treatment outcome would suggest that targeting this locus with TMS could enhance the strength of the protective circuit and/or reduce the risk circuit strength, re-regulating the functional circuits towards configurations associated with longer abstinence. In contrast, the predictive ROI-2 and -3 are on the right hemisphere DLPFC. Although the majority of published neuromodulation studies have targeted the left DLPFC, there are reported anti-craving effects from right hemisphere DLPFC rTMS in various SUDs, including cocaine and alcohol (Camprodon *et al*., 2007; Mishra *et al*., 2010).

Choosing the optimal TMS stimulation location may be the most crucial determinant for TMS treatment effectiveness. However, the parameter space (i.e. location, pulse width, intensity, frequency, etc) to determine optimal neuromodulation treatment is huge to the point of being virtually impossible to fully explore in human clinical studies. Investigators and addiction treatment clinicians have utilized various TMS loci, e.g., F3 from the 10-20 EEG system (Liu *et al*., 2017), or various coordinates with the aid of a neuronavigation system (Pripfl *et al*., 2014; Terraneo *et al*., 2016). Unfortunately, the selection of any location usually lacks a compelling justification and is most often following a previously published study. Fox and colleagues have explored the relationship between the functional connectivity from several DLPFC sub-regions and MDD treatment outcome (Fox *et al*., 2012). The wide range of treatment outcomes from targets all considered to be ‘DLPFC’ illustrates the importance of appropriate target selection.

The present results expand our understanding of the role of DLPFC circuits in cocaine dependence and may suggest future neuromodulation intervention approaches by providing three potential DLPFC stimulation targets. Previous studies employing on-line TMS during fMRI acquisition have demonstrated that following single-pulse DLPFC stimulation (although different target locating strategy), downstream brain regions of DLPFC are activated including the ACC, caudate, and VLPFC (Dowdle *et al*., 2018; Vink *et al*., 2018). Given these acute effects of TMS on fMRI signal, it may be possible to enhance the functional connectivity of the protective circuits and/or reduce the functional connectivity of risk circuits, which may be usefully applied in future therapeutic neuromodulation interventions. Moreover, the strength of these circuits, and how they change over the course of a TMS treatment regime, may potentially serve as a prospective biomarker for treatment efficacy.

Notably, the cumulative statistically predictive influence of these three functional DLPFC sub-region circuits suggests that the variance in cocaine relapse explained by these circuits are at least partially distinct, with the left DLPFC ROI-1 likely primarily related to the executive functioning domain and the two right hemisphere DLPFC ROIs to social-emotional and memory domains. These data driven findings independently corroborate the model of multiple networks/circuits disruptions underlying SUD (Volkow *et al*., 2011). Our findings further suggest that treatment efficacy might be enhanced from a multi-site stimulation design that includes more than one of the identified functional circuits.

Finally, our current analysis focused on the relationship between functional circuits of the DLPFC and cocaine relapse. Other functional circuits have previously been identified that predict cocaine relapse, e.g., between the PCC and the hippocampus (Adinoff *et al*., 2015), between the temporal pole and the mPFC (Geng *et al*., 2017), as well as from large-scale networks such as the executive control network (McHugh *et al*., 2017; Yip *et al*., 2019). However, the current analysis was specifically focused on the DLPFC, motivated by its prominent potential and feasibility as a neuromodulation target, and incorporated contemporary statistical rigor which allowed us to identify two functionally distinct circuits, i.e., a protective and risk circuit, for each of the three predictive ROIs. It is worth noting that each pair of circuits are functionally connected to the same DLPFC ROI, and thus care should be taken if utilizing these predictive ROIs as neuromodulation targets since different individuals might not respond to a given TMS intervention in the same way. This may also explain the variation in treatment outcome seen in neuromodulation treatment studies (see (Hanlon *et al*., 2015), Table 1). Future studies should prospectively test the effects of TMS intervention targeting these functional circuits, on an individual basis, to examine how they change across the treatment regimen and their relationship with treatment outcome efficacy.

### Limitations

Despite the novelty and potential treatment implications of our study, several limitations should be considered. Our sample included only five females and future work should address the possibility of gender differences. We did not include alcohol consumption as a covariate in our prediction model despite its large impact on brain structure and function and its well-known comorbidity with cocaine dependence because this information was originally acquired as an ordinal rather than a continuous variable. However, our participants reported on average consuming less than 2 alcohol drinks in the previous two weeks (based on the ordinal variable from 0 to 6), suggesting a low alcohol influence in the present cohort. Finally, our data were measured after psychosocial treatment, and therefore cannot speak to whether these altered circuits were pre-existing or consequence of the treatment, but do illustrate an important relationship to successful abstinence.

## Conclusions

We built Cox-regression based prediction models using the functional connectivity from 98 surface loci covering the entire DLPFC and identified three loci with high prediction power for cocaine relapse, one on the left hemisphere, which is essentially the same locus that showed TMS treatment efficacy in a previous study, and two on the right hemisphere. We also identified three sets of functionally distinct DLPFC-based circuits related to these ROIs that conveyed protection against and risk for cocaine relapse. These findings support important roles for “bottom up” drive to use drug and “top down” cognitive control over behavior, emotional and social functioning, and dysregulated memory and learning related circuits, consistent with modern models of SUD (Koob and Volkow, 2016; Volkow *et al*., 2011). Future studies need to assess the ability of TMS to alter these circuits in therapeutically useful ways. Finally, these data highlight the importance of the balance between protective and risk factors in determining treatment outcome and the potential utility of resting-state functional connectivity as a clinically relevant biomarker to guide individualized therapy for drug dependence.

## Supporting information

STROBE

## Acknowledgments

The authors would like to acknowledge the Substance Abuse Team at the VA North Texas Health Care System, Homeward Bound, Inc. for their assistance, and Nexus Recovery Center for their support in the screening and recruitment of study participants.

## Supplementary material

Supplementary material is available online.

### Abbreviations

ACC: anterior cingulate cortex
AC-PC: anterior commissure-posterior commissure
AUC: Area-Under-Curve
AUD: alcohol use disorder
BOLD: blood-oxygen-level-dependent
CPD: cigarette per day
CUD: cocaine use disorder
DLPFC: dorsolateral prefrontal cortex
DMN: default mode network
DMPFC: dorsomedial prefrontal cortex
DSM: the Diagnostic and Statistical Manual of Mental Disorders
ECN: executive control network
FFA: fusiform face area
fMRI: functional MRI
IFG: inferior frontal gyrus
IPL: inferior parietal lobule
IQ: intelligence quotient
MCL: mesocorticolimbic
MDD: major depressive disorder
MNI: Montreal Neurological Institute
mPFC: medial prefrontal cortex
MTL: medial temporal lobe
PCC: posterior cingulate cortex
ROC: receiver operating characteristic
ROI: region-of-interest
rTMS: repetitive TMS
SD: standard deviation
SUD: substance use disorders
TE: echo time
TMS: transcranial magnetic stimulation
ToM: theory of mind
TR: repetition time
UDS: urine drug screen
VLPFC: ventrolateral prefrontal cortex
vmOFC: ventromedial orbitofrontal cortex
vmPFC: ventromedial prefrontal cortex

