## supplementary for "Functional connectivity of dorsolateral prefrontal cortex predicts cocaine relapse"

**Content list:**

**Methods.** Detailed analytical pipeline

**Supplementary Fig. 1.** Schematic diagram of analytical pipeline

**Supplementary Table 1.** Brain regions in the group level heat map of protective circuit and risk circuit for predictive ROI-1, predictive ROI-2, and predictive ROI-3

**Methods.** Detailed analytical pipeline

*1. Image preprocessing*

Imaging data preprocessing and functional connectivity analyses were conducted using AFNI (v17.0.06, <http://afni.nimh.nih.gov/afni/>) and SPM12 (<http://www.fil.ion.ucl.ac.uk/spm/software/spm12/>) software packages. The Cox regression analysis was conducted using a customized program on the Matlab platform (R2017a, The MathWorks, Inc., Natick, Massachusetts). Data preprocessing included: discarding the first 5 volumes to allow the magnetic resonance signal to reach steady state, slice timing correction (*3dTshift, AFNI*), volume registration (*3dvolreg, AFNI*), quadratic detrending (*3dDetrend, AFNI*) and head motion correction (*3dTproject, AFNI*). Head motion was also evaluated at the frame-by-frame level to further control quality by using pair-wise displacement calculated based on the Euclidean distance (*1d_tool.py, AFNI*). Volumes with displacement > 0.35 mm were censored, participants were excluded if their mean head motion across volumes were greater than 0.2mm or their percentage of censored volumes exceeding 20%. Two participants were excluded due to head motion exceeding threshold, leaving 43 participants in the final analyses. Signal from white matter and cerebrospinal fluid (CSF) was regressed out as a marker of non-neuronal noise (*3dTproject, AFNI*). A band-pass filter was applied to select low-frequency fluctuations between 0.012Hz and 0.1 Hz (*3dTproject, AFNI*) (1). The blood-oxygen-level-dependent (BOLD) data were normalized to standard MNI image space and resampled to 2mm isotropic resolution (SPM12).

*2. Dorsolateral prefrontal cortex functional connectivity (DLPFC FC)*

As the DLPFC is a large and heterogeneous region, we performed an extensive and systematic analysis of the entire DLPFC (both left and right hemispheres). We first defined the DLPFC borders based on the probabilistic “Harvard-Oxford cortical and subcortical structural atlases” (3^rd^ component, which contains part of superior frontal gyrus, middle frontal gyrus, and inferior frontal gyrus) provided within FSL (v5.0.9, <https://fsl.fmrib.ox.ac.uk/fsl/fslwiki)>. In consideration of both computational efficiency and the stimulation focality of transcranial magnetic stimulation (TMS), for which we intend to provide guidance with our results, we down-sampled this DLPFC mask to 8mm isotropic resolution and selected regions of interest (ROIs) near the surface of the cortex (accessible directly to TMS). This yielded 98 sets of DLPFC coordinates around which we created 98 4mm radius spherical seed ROIs that evenly sampled the surface level of both hemispheres. Cross-correlation coefficient (CC) maps of each participant were generated by correlating the time course of each of the 98 seeds with that of each voxel in the whole brain. Fisher’s Z-transformation was applied to the CC maps resulting in z maps that were used in a subsequent voxel-wise Cox regression. All subsequent analyses were conducted within voxels in a grey matter constrained probabilistic mask.

*3. Voxel-wise Cox regression analysis*

To investigate the relationship between DLPFC FC and cocaine relapse, we utilized the Cox regression model to perform a voxel-wise whole brain search for DLPFC circuits that predicted cocaine relapse, with the factors of age, gender, years of education, daily cigarette use and head movement during scanning as covariates. The beta coefficient (weighting of the model fit) of each voxel (i.e., its connectivity with that DLPFC seed voxel) was estimated via Cox regression. We then obtained the relative hazard ratio (HR) values by calculating the exponential of the beta coefficient values to generate HR maps of all participants.

*4. Thresholding and composite index generation*

All HR maps from the voxel-wise Cox regression were thresholded at *p*<0.001 (for both positive and negative beta coefficients). These HR maps were used to generate: 1) A set of ‘protective’ circuits, which are comprised of voxels with HR value greater than 1 (or positive beta coefficient, indicating higher likelihood of relapse within the follow-up period with stronger FC); and a set of ‘risk circuits’, which are comprised of voxels with HR value less than 1 (or negative beta coefficient, indicating less likelihood of relapse within the follow-up period with stronger FC); and 2) Two composite indices: protective (indexP) and risk (indexR) indices using linear summation of all voxels within the ‘protective’ and the ‘risk’ circuits, respectively, for each participant.

*5. Brain-Behavior relationship (model fitting, ROC analysis and model comparison)*

We fit both indexP and indexR into the final Cox model, with age, gender, years of education, daily cigarette use and head movement during scanning as covariates. A Receiver-Operating-Characteristic (ROC) analysis was conducted to determine the predictive power of the model. The ROC curve was generated by calculating the sensitivity and specificity at multiple thresholds, then the Area-Under-Curve (AUC) was calculated and used as a measure of prediction accuracy. We used the log-likelihood tests for different model comparison.

*6. Cross validation and permutation test*

To validate the relapse prediction models and to test their potential in predicting new individuals, we conducted the above analyses from step 3 to 5 in a leave-one-out (LOO) manner, i.e., we repeated the whole analysis 43 times, excluding one participant each time and using the remaining 42 participants to estimate indices or intermediate results of this individual participant. After all LOO steps were finished, we stacked the HR maps of all 43 LOO steps, binarized and thresholded at 85% to generate the group level heat map, identifying a set of risk circuits and a set of protective circuits from each DLPFC seed that uniquely contributed to cocaine relapse.

Permutation testing was performed to empirically determine significance thresholds and thus to control for overfitting. We repeated the entire analysis 10,000 times, each time with the predictor (indices based on DLPFC FC) and outcome (days till relapse) pairs randomly permuted to generate our data/model specific empirical *null* distribution. The *p*-value of the AUC was then derived based on the ranking of the actual AUC value in this empirical *null* distribution. For our 98 DLPFC ROIs, the AUC values were considered statistically significant if the *p*-values were less than 5.1x10^-4^ (0.05/98 for multiple comparison correction).

**Supplementary Fig. 1.** Schematic diagram of analytical pipeline

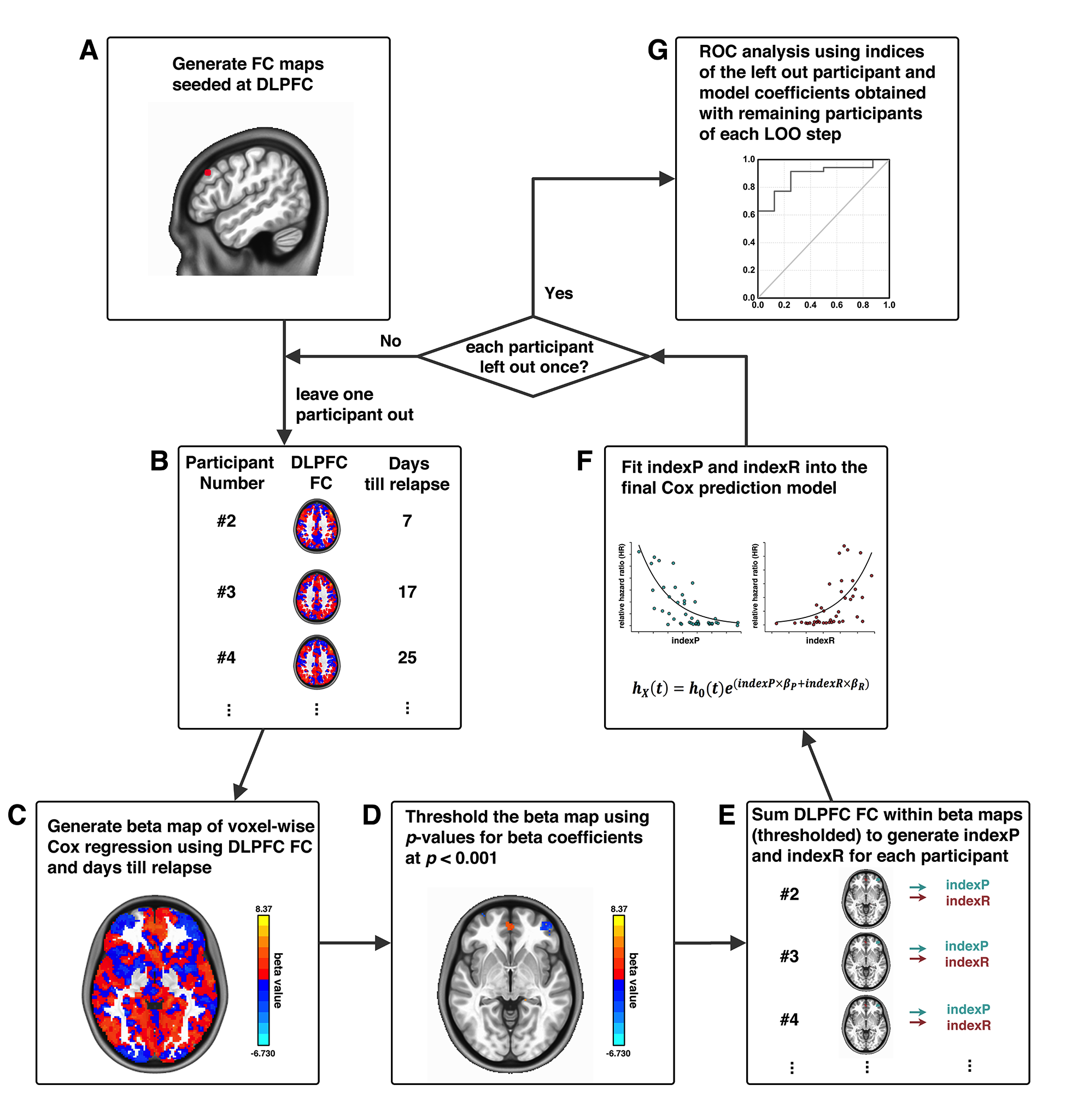

Example of the analytical pipeline from one DLPFC locus. A DLPFC ROI is selected as a seed (A); the whole-brain FC of this DLPFC seed is calculated for each participant (B); a voxel-wise Cox regression is conducted using DLPFC FC and days until relapse to generate beta maps (C); beta maps of Cox regression is thresholded (D); generation of indexP and indexR by linearly summation of the DLPFC FC within the thresholded beta maps for the negative and positive beta voxels, respectively (E); construction of the final prediction model by fitting indexP and indexR into the Cox model (F); the steps from B to F were repeated in a leave-one-out (LOO) manner, and after each participant is left out once, a final ROC analysis evaluates the prediction model (G).

**Supplementary Table 1.** Brain regions in the group level heat map of protective circuit and risk circuit for predictive ROI-1, predictive ROI-2, and predictive ROI-3

| Seed | Circuit | Brain Region | MNI Coordinates (LPI) | | | Cluster Size (mm^3^) |
| --- | --- | --- | --- | --- | --- | --- |
|  |  |  | X | Y | Z |  |
| Predictive ROI-1 | Protective | L-IFG | -44 | 52 | 6 | 600 |
|  |  | L-IPL | -54 | -42 | 54 | 160 |
|  |  | R-IFG | 50 | 46 | -12 | 112 |
|  |  | L-DLPFC | -38 | 36 | 38 | 112 |
|  |  | R-DLPFC | 42 | 22 | 18 | 88 |
|  |  | L-DLPFC | -42 | 10 | 58 | 72 |
|  |  | L-ITG | -58 | -36 | -24 | 56 |
|  |  | L-Insula | -38 | 0 | -2 | 56 |
|  |  | R-DLPFC | 30 | 54 | 8 | 48 |
|  |  | R-IPL | 50 | -50 | 32 | 32 |
|  | Risk | L-Cuneus | -14 | -78 | 38 | 1152 |
|  |  | R-Cuneus | 20 | -66 | 28 | 736 |
|  |  | R-FFA | 36 | -50 | -18 | 632 |
|  |  | B-vmPFC | 0 | 48 | -6 | 336 |
|  |  | R-PostCG | 62 | -24 | 22 | 280 |
|  |  | B-vmPFC | 2 | 44 | -16 | 232 |
|  |  | R-mOFC | 10 | 26 | -26 | 168 |
|  |  | R-Cuneus | 14 | -82 | 46 | 120 |
|  |  | R-Fusiform | 28 | -68 | -18 | 112 |
|  |  | R-PCC | 2 | -42 | 18 | 112 |
|  |  | L-Lingual | -18 | -64 | 4 | 96 |
|  |  | L-Lingual | -20 | -70 | -10 | 88 |
|  |  | L-IOG | -32 | -78 | -10 | 80 |
|  |  | L-Precuneus | -14 | -48 | 34 | 72 |
|  |  | R-Fusiform | 36 | -40 | -14 | 64 |
|  |  | L-Insula | -32 | 24 | 8 | 64 |
|  |  | R-Fusiform | 28 | -82 | -10 | 56 |
|  |  | B-SubCG | 0 | 22 | -12 | 40 |
|  |  | R-MTG | 50 | -42 | 10 | 32 |
|  |  | L-MOG | -32 | -90 | 16 | 32 |
|  |  | R-Precuneus | 8 | -66 | 50 | 32 |
|  |  | L-Thalamus | -2 | -10 | 6 | 24 |
| Predictive ROI-2 | Protective | R-DMPFC | 6 | 50 | 38 | 128 |
|  |  | L-VLPFC | -16 | 36 | -24 | 96 |
|  |  | L-DLPFC | -50 | 18 | 40 | 96 |
|  |  | R-DLPFC | 34 | 46 | 24 | 88 |
|  |  | L-IFG | -50 | 42 | -4 | 72 |
|  |  | L-DLPFC | -40 | 44 | 20 | 56 |
|  |  | L-IFG | -44 | 56 | -10 | 40 |
|  |  | R-IFG | 44 | 50 | 4 | 16 |
|  |  | R-IPL | 44 | -24 | 58 | 16 |
|  | Risk | B-Precuneus | -2 | -50 | 62 | 2096 |
|  |  | L-MTG | -48 | -70 | 2 | 176 |
|  |  | R-Fusiform | 30 | -60 | -18 | 128 |
|  |  | R-PreCG | 48 | 6 | 24 | 128 |
|  |  | L-SMA | -10 | -8 | 64 | 120 |
|  |  | L-SMA | -4 | -8 | 52 | 112 |
|  |  | R-MTG | 44 | -58 | 22 | 80 |
|  |  | L-Thalamus | -6 | -12 | 14 | 64 |
|  |  | L-Hippo | -34 | -22 | -16 | 56 |
|  |  | R-Lingual | 4 | -70 | 2 | 56 |
|  |  | L-Precuneus | -4 | -76 | 46 | 56 |
|  |  | L-Precuneus | -14 | -42 | 46 | 56 |
|  |  | L-STG | -50 | -36 | 16 | 48 |
|  |  | R-Amy | 22 | 0 | -20 | 40 |
|  |  | R-STG | 56 | -30 | 14 | 40 |
|  |  | R-MOG | 50 | -80 | 18 | 40 |
|  |  | L-Amy | -28 | -4 | -18 | 32 |
| Predictive ROI-3 | Protective | R-DLPFC | 34 | 46 | 24 | 200 |
|  |  | L-DMPFC | -8 | 6 | 56 | 136 |
|  |  | R-DLPFC | 46 | 42 | 20 | 72 |
|  |  | L-DLPFC | -26 | 50 | 28 | 48 |
|  |  | R-SFG | 26 | 56 | 4 | 40 |
|  |  | R-Precuneus | 14 | -48 | 40 | 40 |
|  |  | R-DMPFC | 4 | 44 | 44 | 24 |
|  | Risk | L-Precuneus | -16 | -70 | 24 | 1208 |
|  |  | R-Precuneus | 24 | -62 | 20 | 760 |
|  |  | L-Cuneus | -8 | -64 | 10 | 632 |
|  |  | R-Cuneus | 10 | -56 | 6 | 504 |
|  |  | L-MTG | -42 | -88 | 18 | 448 |
|  |  | L-Fusiform | -30 | -46 | -20 | 440 |
|  |  | L-Precuneus | -4 | -48 | 58 | 200 |
|  |  | L-Fusiform | 26 | -46 | -20 | 176 |
|  |  | R-Fusiform | -24 | -52 | -4 | 136 |
|  |  | R-IFG | 44 | 32 | 12 | 104 |
|  |  | R-MTG | 46 | -76 | 18 | 88 |
|  |  | R-SOG | 24 | -66 | 34 | 88 |
|  |  | R-mOFC | 6 | 22 | -28 | 64 |
|  |  | L-Fusiform | -30 | -60 | -10 | 56 |
|  |  | L-MFG | -24 | 28 | -16 | 40 |
|  |  | L-Thalamus | -8 | -28 | 10 | 32 |
|  |  | R-Parahippo | 20 | -18 | -20 | 16 |

*Abbreviations:* IFG, inferior frontal gyrus; IPL, inferior parietal lobule; DLPFC, dorsolateral prefrontal cortex; ITG, inferior temporal gyrus; FFA, fusiform face area; vmPFC, ventromedial prefrontal cortex; Post CG, postcentral gyrus; mOFC, medial orbitofrontal cortex; PCC, posterior cingulate cortex; IOG, inferior occipital gyrus; SubCG, subcallosal gyrus; MTG, middle temporal gyrus; MOG, middle occipital gyrus; DMPFC, dorsomedial prefrontal cortex; VLPFC, ventrolateral prefrontal cortex; Pre CG, precentral gyrus; Hippo, hippocampus; STG, superior temporal gyrus; Amy, amygdala; SFG, superior frontal gyrus; SOG, superior occipital gyrus; MFG, middle frontal gyrus; Parahippo, parahippocampal gyrus.
